# Detailed Overview of the Buildout and Integration of an Automated High-Throughput CLIA Laboratory for SARS-CoV-2 Testing on a Large Urban Campus

**DOI:** 10.1101/2021.09.13.21263214

**Authors:** Lena Landaverde, David McIntyre, James Robson, Dany Fu, Luis Ortiz, Rita Chen, Samuel M.D. Oliveira, Andy Fan, Amy Barrett, Stephen P. Burgay, Stephen Choate, David Corbett, Lynn Doucette-Stamm, Kevin Gonzales, Davidson H. Hamer, Lilly Huang, Shari Huval, Christopher Knight, Diane Lindquist, Kelly Lockard, Trevor L. Macdowell, Elizabeth Mauro, Colleen McGinty, Candice Miller, Maura Monahan, Randall Moore, Judy Platt, Jeffrey Roy, Tracey Schroeder, Dean R. Tolan, Ann Zaia, Robert A. Brown, Gloria Waters, Douglas Densmore, Catherine M. Klapperich

**Affiliations:** Precision Diagnostics Center, Boston University, Boston, MA; Biomedical Engineering, College of Engineering, Boston University, Boston, MA; Boston University Clinical Testing Laboratory, Boston University, Boston, MA; Design, Automation, Manufacturing, and Prototyping Laboratory, Boston University, Boston, MA; Biological Design Center, Boston University, Boston, MA; Software & Application Innovation Lab, Hariri Institute for Computing, Boston University, Boston, MA; Department of Biology, Boston University, Boston, MA; Electrical and Computer Engineering, Boston University, Boston, MA; Office of the Provost, Boston University, Boston, MA; Office of External Affairs, Boston University, Boston, MA; Information Services & Technology, Boston University, Boston, MA; Health Information Privacy Resources, Boston University, Boston, MA; BU Healthway, Boston University, Boston, MA; Office of Research, Boston University, Boston, MA; Department of Global Health, Boston University School of Public Health, Boston University, Boston, MA; Section of Infectious Disease, Department of Medicine, Boston University School of Medicine, Boston, MA; National Emerging Infectious Disease Laboratory, Boston, MA; Office of General Counsel, Boston University, Boston, MA; Health Privacy and Compliance, Boston University, Boston, MA; Continuous Improvement & Data Analytics, Boston University, Boston, MA; Operations, Boston University, Boston, MA; Sourcing and Procurement, Boston University, Boston, MA; Student Health Services, Healthway, Boston University School of Public Health, Boston, MA; HFI Laboratory, Boston University, Boston, MA; Occupational Health Center, Boston University, Boston, MA; College of Engineering, Boston University, Boston, MA; Office of the President, Boston University, Boston, MA; College of Health and Rehabilitation Services, Sargent College, Boston University, Boston, MA

## Abstract

In 2019, the first cases of SARS-CoV-2 were detected in Wuhan, China, and by early 2020 the cases were identified in the United States. SARS-CoV-2 infections increased in the US causing many states to implement stay-at-home orders and additional safety precautions to mitigate potential outbreaks. As policies changed throughout the pandemic and restrictions lifted, there was an increase in demand for Covid-19 testing which was costly, difficult to obtain, or had long turn-around times. Some academic institutions, including Boston University, created an on-campus Covid-19 screening protocol as part of planning for the safe return of students, faculty, and staff to campus with the option for in-person classes. At BU, we stood up an automated high-throughput clinical testing lab with the capacity to run 45,000 individual tests weekly by fall of 2020, with a purpose-built clinical testing laboratory, a multiplexed RT-PCR test, robotic instrumentation, and trained CLIA certified staff. There were challenges to overcome, including the supply chain issues for PPE testing materials, and equipment that were in high demand. The Boston University Clinical Testing Laboratory was operational at the start of the fall 2020 academic year. The lab performed over 1 million SARS-CoV-2 RT-PCR tests during the 2020-2021 academic year.

## Introduction

### Impact of Covid-19 in Boston and Boston University

In late 2019, severe acute respiratory syndrome coronavirus 2 (SARS-CoV-2), a novel coronavirus, was first reported in Wuhan, China^1–3^. Cases in the United States were documented in Washington State on January 20, 2020, and shortly after the World Health Organization (WHO) declared the Coronavirus disease 2019 (Covid-19) pandemic in March of 2020^4,5^. At the height of the SARS-CoV-2 pandemic, individuals of all occupations were taking additional precautions during stay-at-home orders to ensure public safety and health. As the demand for testing increased in parallel with state restrictions lifting and increasing cases in the US, Covid-19 screening was either unavailable, costly, or had test-to-result times too long to work as a screening tool^6^. As part of the initial shutdowns in March 2020, Boston University (BU) went remote and finished the remainder of the semester with online course work. At that time, we began to plan for the return of students in August 2020 which included the construction of a new high-throughput testing laboratory to maintain a testing cadence and turnaround time sufficient to control viral spread on campus^7^.

For BU, SARS-CoV-2 screening testing was part of a multi-faceted strategy to permit in person teaching in the fall of 2020. There were examples of newly formed testing facilities with the same purpose, one of the first and most notable was a team at University of California Berkeley that provided a detailed blueprint for converting the Innovative Genomics Institute to test for SARS-CoV-2 in the university and local community^8^. In Europe, the Francis Crick Institute developed the Crick Covid-19 Consortium with publicly available standard operating procedures (SOP)^9^ for other organizations to follow. At Boston Medical Center, the Center for Regenerative Medicine extended the capacity of the BMC Department of Pathology and Laboratory Medicine to perform RT-qPCR Covid-19 testing with a 24-hour turn-around time^10,11^. These successes led us to explore doing the same for our entire campus, including 45,000 faculty, students, and staff^12^. A team was quickly assembled to stand up an on-site high-throughput clinical laboratory from the ground up. The goal was to enable faculty, staff, and students to safely return with an option for in-person classes, a program known as BU’s Learn from Anywhere (LfA)^7^ during the 2020-2021 academic year. Across the US, many institutions were making similar plans and it is also important to credit the plethora of online collaboration and communication platforms like Slack where scientists from academic, government, and industry came together to assist each other in these endeavors.

By Fall of 2020, BU implemented a multi-stage plan to perform screening testing of approximately 45,000 students, faculty, and staff for Covid-19. The BU campus comprises three locations in Boston and Brookline, MA. The largest location is the Charles River Campus (CRC), which is approximately three miles from the next largest Boston University Medical Campus, followed by the smaller, Fenway Campus. The proposed plan included an on-site testing facility, collection sites, and contact tracing^7^. Instrumental to the plan was building a clinical testing facility with the capacity to test students, staff, and faculty weekly. The development of the Boston University Clinical Testing Laboratory (BU CTL) was a collaborative effort between the BU Office of Research, the Precision Diagnostics Center (PDC), and the Design, Automation, Manufacturing, and Prototyping (DAMP) Laboratory. Specifically, the development and implementation of the facility required a combination of automation, assay development and systems engineering and management. In addition, the new BU CTL would have to meet regulatory and legal requirements set forth under the Clinical Laboratory Improvements Amendments (CLIA) and Massachusetts state law as well as apply for a Food and Drug Administration (FDA) Emergency Use Authorization (EUA) for a laboratory developed test for Covid-19^13^.

### Major decision points

The BU CTL stands out as a technically advanced, purpose built, high throughput automated testing facility. We implemented a high sensitivity RT-qPCR test, integrated automation to support the required throughput, and developed a customized Laboratory Information Management System (LIMS) infrastructure. The sample preparation and RT-qPCR assay were developed to meet both EUA and CLIA requirements (Fig. 1).

**Figure 1.**
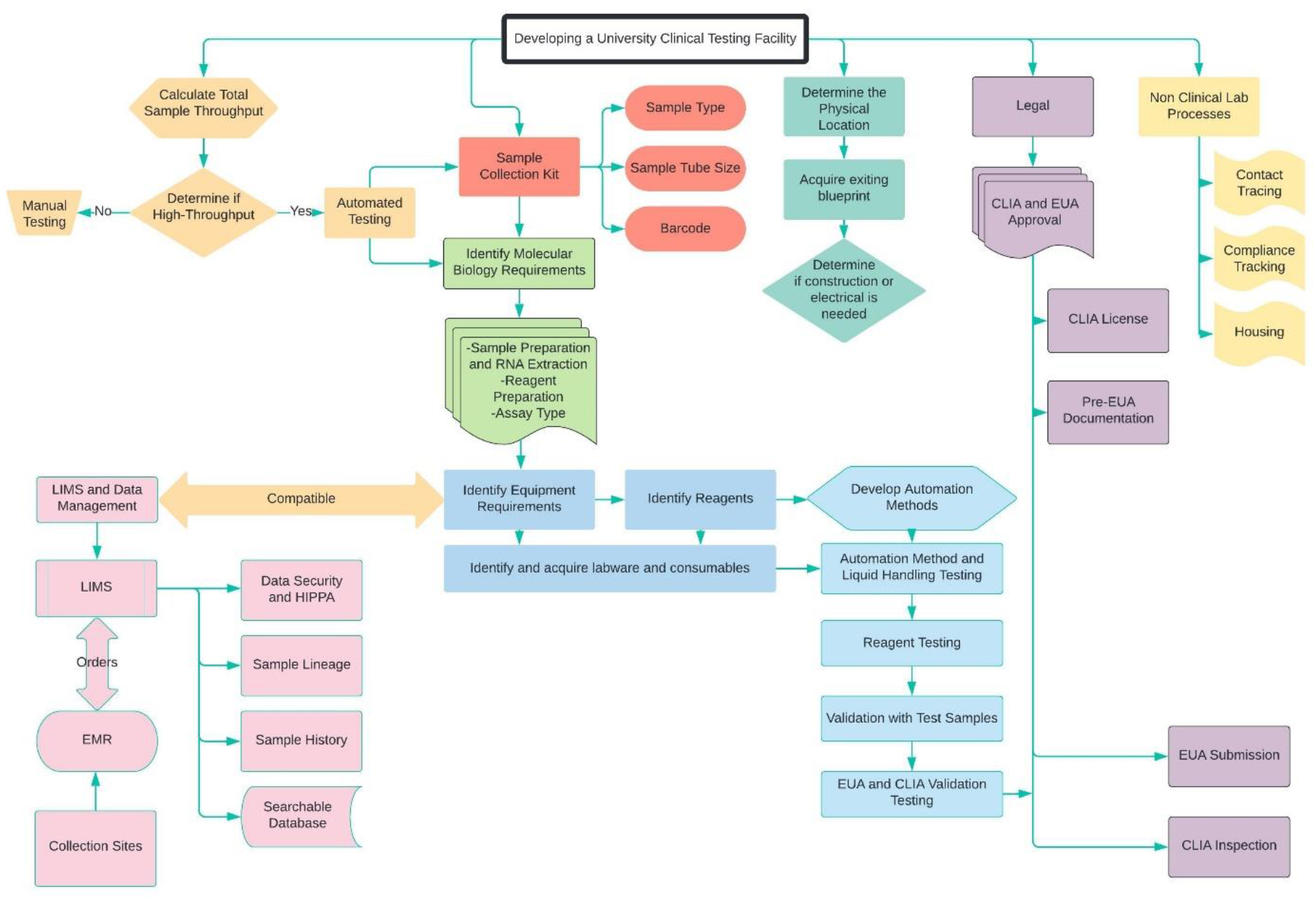
The multi-staged approach for building the Boston University Clinical Testing Laboratory. Each branch builds upon strategic decisions made with the best available information at the time and critical to the function of the CTL. Each branch breaks down different categories to demonstrate how each connect to the physical laboratory. Included here are also considerations made that are tangential to the build out of the automated testing process.

After reviewing various predictive models of SARS-CoV-2 transmission on campus, BU decided to test undergraduate students twice a week and all others based on an assigned testing category with routine asymptomatic screening^7^. This resulted in a projected test load of 5,000 tests/day with a required next day turnaround time. Building the physical and digital infrastructure required a university-wide team to source materials, equipment, and human resources to plan and build the space. The team comprised of Sourcing and Procurement, Office of Research, Legal, Medical Advisory Board, Marketing and Communications, is shown in Figure 2. The project was driven by the following design requirements: adequate laboratory space, safe and approved sample collection and transportation, efficient RNA purification, assay development, automation, sample lineage tracking, CLIA, EUA, and staffing. An ongoing challenge was equipment, material, and PPE supply chain issues caused by the global pandemic^14^. Availability of resources and equipment was a major driving factor in the decisions (Fig. 1) made to build the high throughput clinical laboratory.

**Figure 2.**
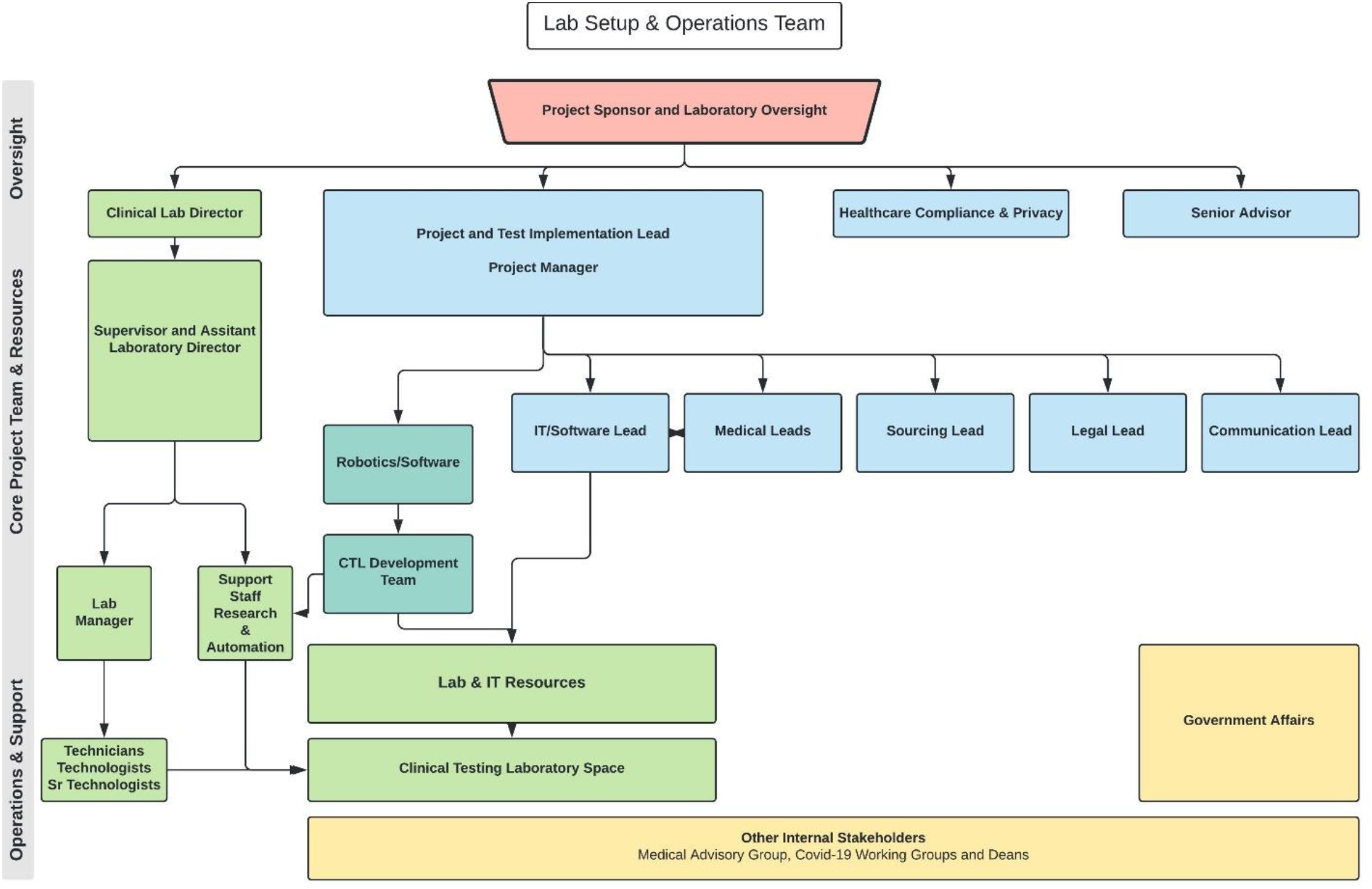
BU’s CTL buildout was a multi-departmental effort within the university and a collaboration of both the Charles River Campus and Boston University Medical Campus. The groups involved were integral to developing many of the important campus support of Covid-19 screening testing including contact tracing and housing.

### Planning and implementation

#### Identifying a space for CTL for high throughput automated testing

Identifying a dedicated space on BU’s CRC was critical. It needed to house sample receiving, automation robots, qPCR machines, and all auxiliary equipment with the appropriate laboratory footprint. A laboratory space was identified within the Rajen Kilachand Center for Integrated Life Sciences & Engineering. The space was initially designed to house yet to be purchased automation equipment for the Design Automation Manufacturing Prototyping (DAMP) Laboratory, so many of the basic infrastructure needs were in place.

Although the space had some of the required infrastructure, additional renovations were necessary to convert the space into a clinical testing laboratory. The CTL layout required separated stations to mitigate contamination, maintain order, and follow the streamlined workflow illustrated in Figure 3. Samples follow a defined workflow upon arrival in the CTL to maintain sample lineage and tracking with each station defined by the process performed. Electrical work included installing additional emergency power outlets to support critical instrument, refrigeration, and freezer units. Additional ceiling support was added for uninterrupted power supplies (UPS) as backup for the critical robotics and qPCR machines. The space was physically modified with doors to include separated entrances for gowning and to close off the initially open, shared space; all doors were secured by key or swipe access only. This included an additional adjacent space incorporated to house additional refrigeration, freezers, and the maintenance of a small existing CLIA testing facility.

**Figure 3.**
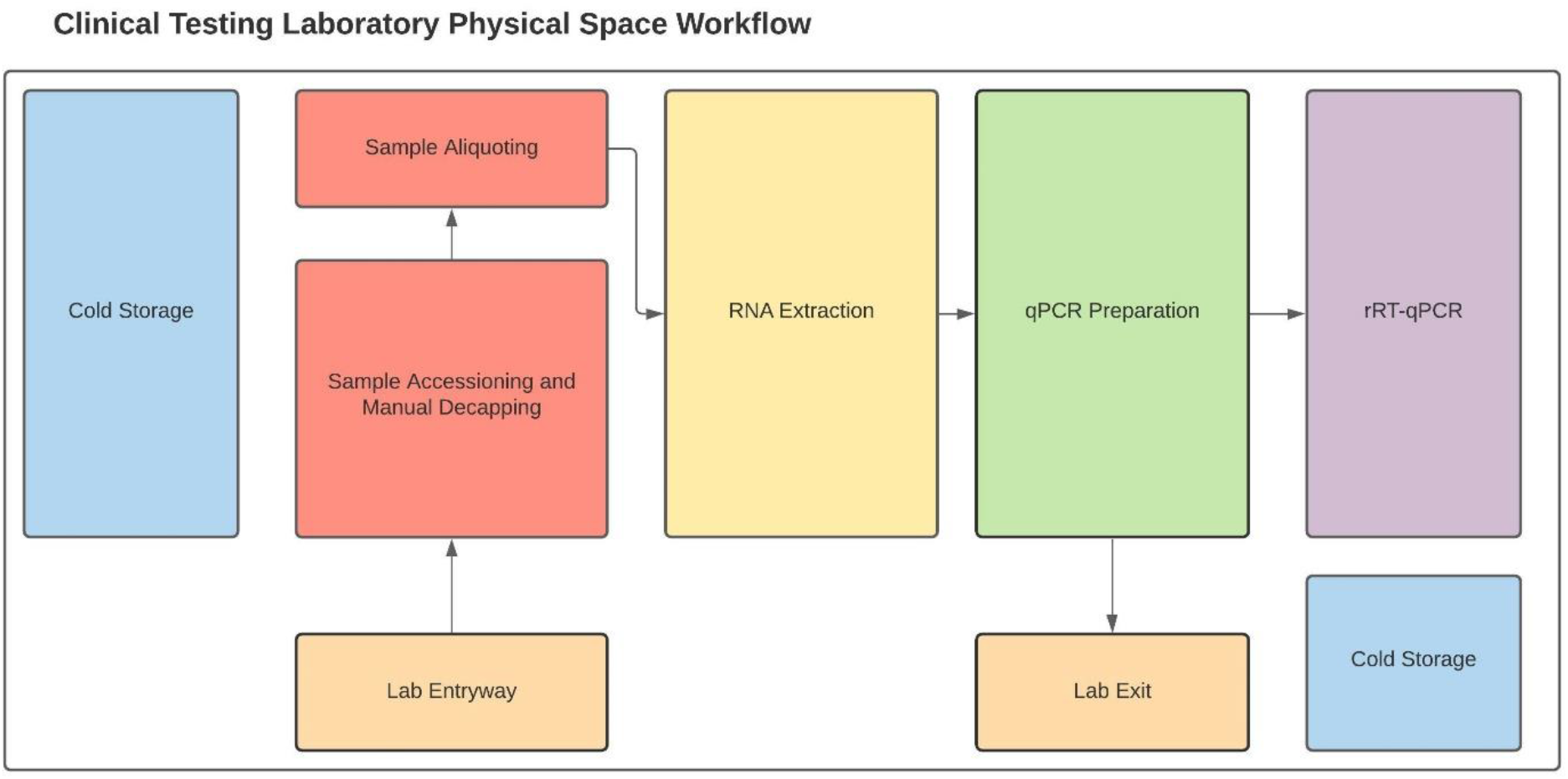
The CTL workspace was divided into sections based on the tasks performed in each section: Sample Accessioning, Sample Aliquoting, RNA Extraction, qPCR Preparation, and RT-qPCR. Specific considerations were made to minimize cross contamination and to isolate the qPCR preparation station away from other processes. Each laboratory staff member would gown and wear appropriate PPE upon entry. Also included in the diagram is the dedicated cold storage spaces for reagents and samples.

#### Sample collection sites

Anterior nares (lower nostril) samples were self-collected under observation. BU set up five sample collection sites: four on the CRC campus and one on the BUMC campus. Remote observed collections are also performed for individuals in quarantine^7^. All students, faculty and staff are required to complete a daily symptom attestation before coming to campus. Asymptomatic individuals receive an electronic clearance badge that they must present upon entering the test site. After check-in they sanitize their hands and approach a check-in station. Everyone is handed a sample tube with a unique barcode and moves to a swabbing station. The observer provides the swab at the swabbing station. Observed swabbing occurs in large, windowed cubicles to allow for social distancing while the individual removes their mask to swab. A time study showed that the entire process of arriving at the site, checking in, and swabbing requires less than 5 mins for almost all users.

All sample collection sites have windowed check-in and self-collection booths, clear labeled signs and directional arrows on the floor spaced for social distancing, and sanitizer dispensers available between stations. To ensure a clean, sterile surface between collections, a sealed single swab is placed on top of sheet of parchment. During sample collection the tube cap can be placed upside down on the parchment paper while the uncapped tube is placed in a small metal cup to hold the tube and prevent spilling. The parchment paper, swab wrapper, and broken off end of swab were disposed of in trash receptacles. A Health Insurance Portability and Accountability Act (HIPPA), Hazardous Material Regulations (HMR), and Occupational Safety and Health Administration (OSHA) certified courier transfers samples on a scheduled basis to the CTL. To ensure the safe travel of the samples and compliance with biosafety requirements samples are packaged in Test n’ Toss Disposable Test Tube Racks (Whitney Medical Solutions, Niles, Illinois) that are contained in a sealed bag. The sealed containers are transported in customized corrugated cardboard boxes labeled with biohazard information, return address, and delivery address.

### Development of sample testing process flow

#### SARS-CoV-2 and CDC approved testing methods

In early 2020, the gold standard testing strategy recommended by the Center for Disease Control and Prevention (CDC) was testing the nucleocapsid (N) gene in 3 target regions as a singleplex RT-qPCR^6,15^. The CDC introduced the first emergency use authorized (EUA) primer and probe set^16^. At the time the laboratory was establishing an assay protocol, two targets (N1 and N2 with RNase P (RP) as the human material control) were required to identify positive cases. There were also limited multiplex RT-qPCR options with EUAs for clinical use. An example of a widely available option was the TaqPath Covid-19 combo kit^17^ (Thermo Fisher, Waltham, MA) that targeted the S (spike), ORF1ab, N regions and included a spiked in internal control ms2phage. However, the kit at the time did not contain a human specific control and was costly even at scale for our application^17^.

We designed a laboratory developed real-time reverse transcription polymerase chain reaction (RT-qPCR) test (LDT) and submitted it for FDA EUA in July of 2020. The BU SARS-CoV-2 Test uses primer and probe sets (IDT Custom, Coralville, Iowa) targeting the N1 (FAM-Tagged) and N2 (YAK-Tagged) SARS-CoV-2 genetic sequences and one human cellular material control (RNase P (Cy5-Tagged)) in each well. Overall performance of the assay automation workflow requires incorporation of external run controls on each plate, including a positive template control (2019-nCov, nucleocapsid gene), a negative extraction control (NEC) and a no template control (NTC). The assay uses the CDC EUA kit sequences for the BU-SARS-CoV-2 Test in a multiplexed assay^15^. The costs for the 2020-2021 academic year averaged $12.70 per test.

### Overview of testing method

#### Biosafety and personal protective equipment

A biosafety SOP was developed following CDC guidelines in collaboration with the BU Environmental Health and Safety (EHS). All samples arriving to the laboratory are counted and inventoried by technicians without opening packaging and are placed into a 4°C fridge until further processing. The first processing step is to heat inactivate samples in a dry bead bath under a biosafety hood. All specimens remain closed until after this step. Lab staff wear fresh face masks, disposable lab coats, and gloves while in the lab. When handling active samples, individuals are required to wear an additional back tying disposable lab gown, face shield and booties. Samples follow a unidirectional flow to maintain sample lineage and minimize any chance of cross contamination (Fig. 3).

#### Automation with Hamilton Microlab STAR Robotic Systems

The BU CTL is outfitted with seven configured Hamilton Microlab STAR (Reno, NV) automated liquid handling systems. The instruments run protocols written in Hamilton’s VENUS software with programmable hardware and integrated data handling. The configurability of the Microlab STAR is unique from other all-inclusive or single purpose systems which include instruments such as KingFisher (ThermoFisher), QIAsymphony (Qiagen), or MANTIS (Formulatrix). The Microlab STAR can be modified to execute multiple tasks due to their flexible deck layout and modular components that lends to developing unique integrated automation protocols. Additionally, the instruments can be adapted to various assay and sample preparation protocols when large-scale COVID-19 testing is no longer needed.

Hamilton’s direct from manufacture consumables, equipment, and parts were integral to ensure receipt and installation on schedule for the July 2020 piloting of testing. We considered other systems but ran into supply chain issues for instruments and consumables as the US rapidly began to scale up testing in mid-2020. For example, the ThermoFisher KingFisher instrument was listed as authorized equipment on several EUAs including the ThermoFisher TaqPath EUA, drastically reducing their availability. Lastly, the support provided by the engineers from Hamilton was integral to getting the systems set up and processing samples in 12 weeks. Automation methods were first validated in laboratory with water, reagents, and utilizing well characterized discarded samples from other CLIA testing laboratories.

The sample processing throughput was mapped for the planned 5000 tests per day over a 7-day period by estimating the total run times on instruments including reagent preparation and manual labor (Figure 4). The calculation and estimates were a combination of times for manual labor and automation instrumentation time. Reagent preparation and loading time is accounted for between instrument runs. The final calculation accounting for manual processes is a total of 6200 tests processed per day with next-day results. The entire automation process includes 7 MicroLab STAR systems: 1 for sample aliquoting, 3 for RNA purification and extraction, and 3 for qPCR Preparation (Fig. 4A).

**Figure 4.**
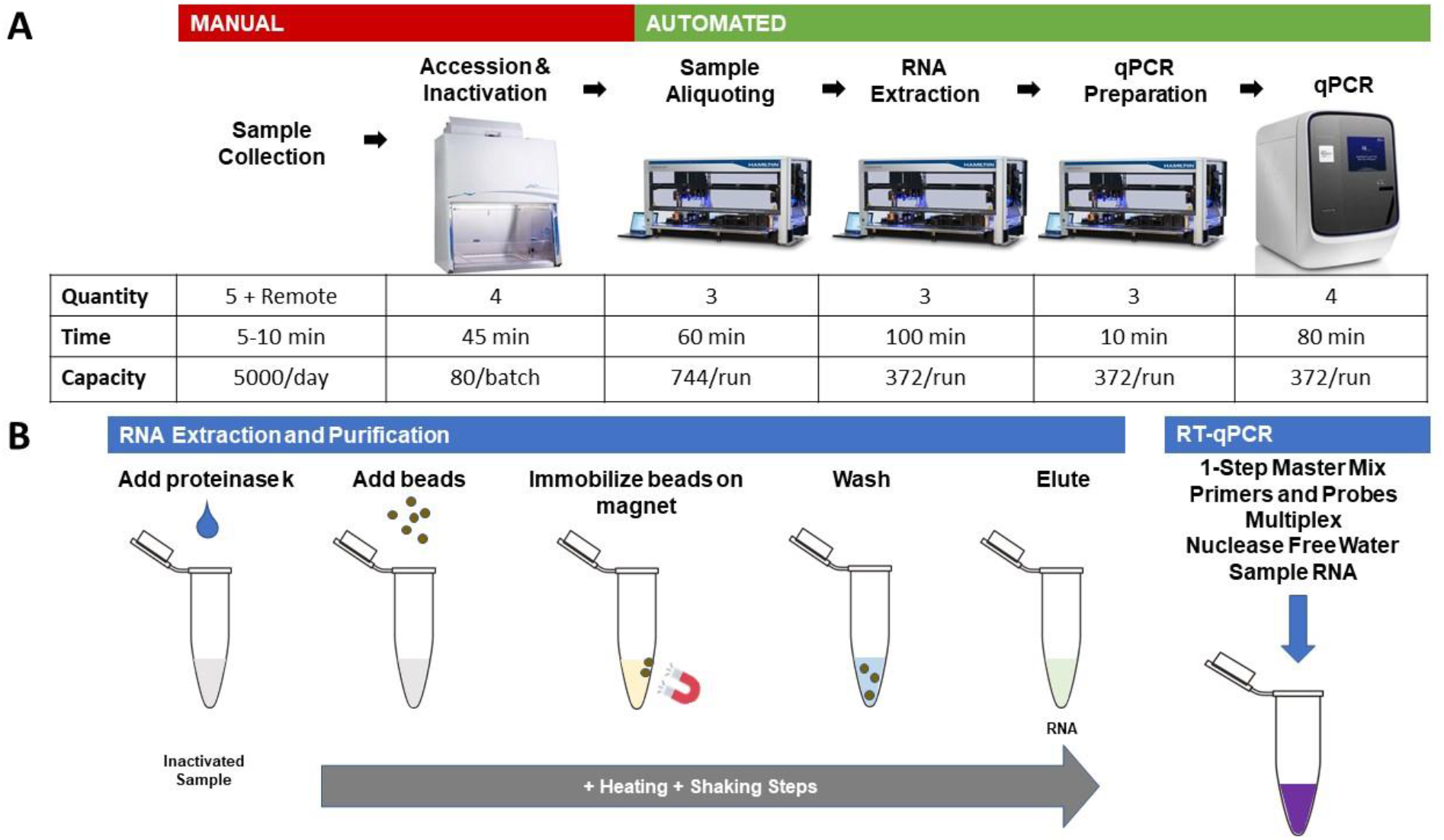
A. The BU CTL workflow begins at the manual step of Accession & Inactivation steps. The automation steps include 3 Microlab STAR Systems with high throughput specific methods for the BU workflow: Sample Aliquoting Microlab STAR, RNA Extraction Microlab STAR, and qPCR Preparation Microlab STAR. The table indicates the time required for each step not including the reagent preparation and loading, cleaning steps, and sample or plate loading. B. The two critical assay steps include RNA Extraction and Purification - using magnetic bead extraction - and RT-qPCR preparation. The two steps were modified from the original protocols to be supported on the Hamilton Microlab STAR in a high-throughput automated workflow.

#### Anterior nares swabs and collection vials

Nasopharyngeal (NP) swabs were the gold standard collection method for Covid-19 testing using RT-qPCR at the time of establishment of the CTL. These tests were typically performed within a hospital or clinic using a process similar to influenza sample collection. However, as the need for SARS-CoV-2 testing increased, there was a worldwide shortage of NP swabs and viral transport media (VTM) used in collection tubes. Detailed investigations were made into alternative sample types, this included saliva, buccal, and anterior nares (AN) collections.

Although each sample collection type had its advantages, the AN swab met multiple requirements. Because they are less complicated to make, AN swabs were more widely available and by late spring of 2020 had been previously EUA approved as a sample type collection type for Covid-19^18^. An additional advantage of AN swabs is that they can be used for self-collection especially with a non-hazardous buffer such as Phosphate Buffer Solution (PBS) or Saline. At the time of assay development, there was minimal information and documentation on the success of saliva sample types for large scale asymptomatic clinical testing. There were known issues with sample viscosity and automation instrumentation, so we selected AN swab.

Due to the high demand of materials, intensive research was performed on identifying compatible collection kits. BU sourced collection kits from Puritan Medical Products and in conjunction with collection kits from other vendors. The finalized selection of collection materials was determined by rigorous testing and comparison of collection tube and swab parameters. Tube size, barcode type, barcode placement, and sample volume were critical for automation compatibility. All sample collection tubes are loaded onto either 24 or 32-tube sample carriers (Hamilton Robotics Company) which can carry either 14.5-18 mm or 11-14 mm outer diameter tubes, respectively. Sample barcodes are read through a window on the sample carriers by the Autoload (Hamilton Robotics Company) and must meet instrument specifications (ML STAR Autoload Specifications). During aspiration steps, it was essential that the robot tips could reach the surface of the sample liquid and reach a minimum depth in the tube to account for volume displacement. Customized 3D printed risers (uPrint SE, Stratasys, Eden Prairie, Minnesota) were also developed to ensure each individual tube was at the correct height. Lastly, multiple AN swab options were tested for swab quality (minimal shedding of material) and swab breakpoints (location the swab would be broken in the tube). Swab breakpoints determined if the swab would be compatible with the height of the tube while also being easily removed by staff before the initial aspiration step. Our vials included ones with combined swab and caps or a patented cap that allows for a swift uncapping and swab removal in one action.

#### Inactivation, accessioning, and sample aliquot

Samples arrive at the laboratory and are first processed manually for inactivation and sample accessioning. The laboratory processes each sample by disinfecting the tubes with 70% ethanol and inactivating samples at 95°C for 10 min. inside a biosafety cabinet. Once inactivated, samples are scanned into the LIMS system and marked as received before being uncapped and loaded onto carriers for the Sample Aliquot Hamilton Microlab STAR. The Sample Aliquot Microlab STAR is a 16-channel system that consists of tip carriers, sample carriers, plate carriers, tube carriers, and an autoload that aspirates and dispenses samples into 96-well deep-well plates for nucleic acid extraction.

### RNA extraction

Automated isolation of nucleic acids from crude sample material was achieved using the MagMAX™ Viral/Pathogen II (MVP II) Nucleic Acid Isolation Kit (ThermoFisher, Waltham, MA). The extraction kits include magnetic beads with a silica coated surface and a magnetic core. Nucleic acids are absorbed to the silica-surface of the beads in the presence of isopropanol and high concentrations of chaotropic salts, which remove water from hydrated molecules in solution. Once bound to the magnetic beads, the nucleic acids can be separated from the solution with a magnet. Polysaccharides and proteins do not adsorb to the beads and are removed by subsequent washing. Pure nucleic acids are then eluted from the beads by applying low-salt conditions and heat (Fig. 4B). The MagMAX™ Kit’s manual protocol^19^ was modified and scaled to be compatible with our automation. These changes included an additional ethanol wash step, dead volumes, and reusing waste aspiration tips. At the time of development, there was no published Hamilton Method for the MagMAX™ Kit that accounted for tip reuse, maximizing the deck layout, and integrated with sample data capturing.

The three RNA Extraction Hamilton Microlab STAR instruments’ deck layouts include tips carriers, tip isolators, reagent carriers, Hamilton Heater Shaker (Hamilton Robotics Company, Reno, NV), and Magnum FLX magnetic ring stand (Alpaqua, Beverly, MA). The system can process a maximum of 4 plates in a single run and takes approximately 2.5 hours. The plate map information from the Sample Aliquot step is mapped to the final elution plates when the RNA Extraction method is complete.

Challenges resulted from unique liquid properties and an effort to reduce tip waste in the protocol. Initial validation with reagents and discarded samples exposed contamination issues from droplets generated during liquid waste disposal in the Hamilton MFX Gravity Waste Module, unwanted liquid retention in tips, and the formation of bubbles post-reagent dispense. The method was tested between liquid class modifications by implementing a checkered Extraction Plate layout with alternating nuclease free water and positive control samples to look for and eliminate cross well contamination. Changing dispense and aspiration speeds within the liquid classes module resolved contamination issues.

### qPCR preparation

To maximize the test throughput and minimize turnaround time, we consolidated 4 elution plates from the extraction robots into one 384-well plate for qPCR. Consolidation is performed by the qPCR Prep Hamilton Microlab STAR instruments. There are 372 purified RNA samples per plate and 9 control wells. Lab Technologists prepare reaction mixes that contain the Applied Biosystems TaqPath Master no ROX Master Mix (ThermoFisher, Waltham, MA), Nuclease Free Water, and IDT Primer and Probe Mix (IDT Custom) (Integrated DNA Technologies, Coralville, IA). The three controls are transferred to the qPCR plate during the PCR set up steps. Each 384 well qPCR plate will contain 1 Positive qPCR Addition Control, 4 NECs and 4 NTCs. A human cellular material control, RNase P (RP), is expected to be present in all valid samples and the NEC. RNase P acts as both an extraction control and an internal control.

The qPCR Prep Hamilton Microlab STAR is used to set up the RT-qPCR reactions. It has the shortest run time of all robots in the process and uses the CO-RE 96-probe Head to quickly aspirate the reaction mix from a deep well midi plate into the 384-well plate followed by addition of the purified RNA samples with mixing. The challenge with this step is the liquid handling for the viscous reaction mix, which can lead to failed RT-qPCR runs. Specific to automation, optimization programming settings for liquid classes for the reaction mix were made by adding a mix step prior to aspirating with no following and no blow out. The positive qPCR control is added manually by the CTL staff after the plate is filled. The plate is then manually sealed and loaded onto the QuantStudio 7 Flex. Output files are created by the qPCR Prep Hamilton Star which are formatted for the QuantStudio 7 Flex. Data from the run is analyzed on the Design and Analysis 2 (Applied Biosystems) software and results are confirmed by a technologist and the Clinical Lab Supervisor.

### Laboratory integration of LIMS

Tracking data and sample lineage is critical to the management of the information produced in the laboratory. The CTL implemented a LIMS that receives orders from the EMR systems, receives data captured from automation instruments, manages sample lineage tracking, and conducts reporting (Fig. 6). An example of a step captured by the LIMS system occurs during accessioning, where each individual sample is updated in the LIMS system to indicate that it has been received in the laboratory. The seamless and paperless capture of all information optimizes the workflow while maintaining data integrity and security. We depend on secure APIs for data transfer between the automation instrumentation and LIMS. To support the EMR reporting and test ordering, HL7 integrations were developed with the two independent EMR systems for faculty and staff, and students.

**Figure 5.**
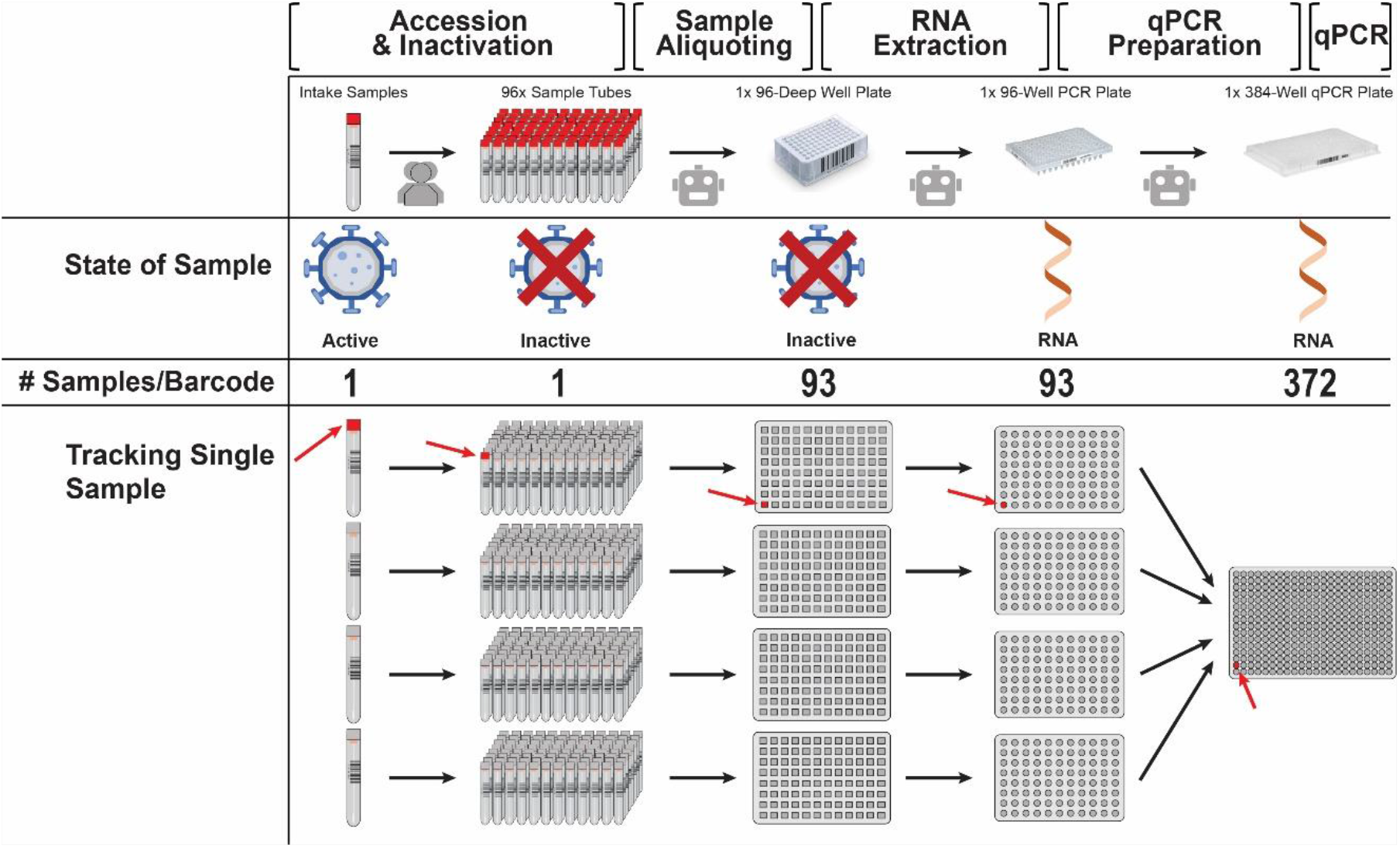
Each sample has a unique barcode ID that is linked to the plate and well location. State of Sample shows the progression from active, to inactivated, and extracted. Each step in the process is automated except for Sample Intake and Accession. The samples are inactivated in the original tubes prior to tube opening. Each qPCR plate contains 372 samples with qPCR and extraction controls. Lab technologists load tubes onto the sample carriers that are pulled in by an Autoload. They manually load the tips, barcoded plates, and extraction controls onto the instrument according to dialog prompts from the Sample Aliquot Method within the Venus Software. The program method associates all the individual samples to the plate and well location. Controls included on each 96 well extraction plate as follows; 1 NEC and 1 NTC. The NEC is composed of pooled discarded negative samples and the NTC is Nuclease Free Water (Integrated DNA Technologies, Coralville, Iowa). The system can aliquot up to 744 samples in one hour.

**Figure 6.**
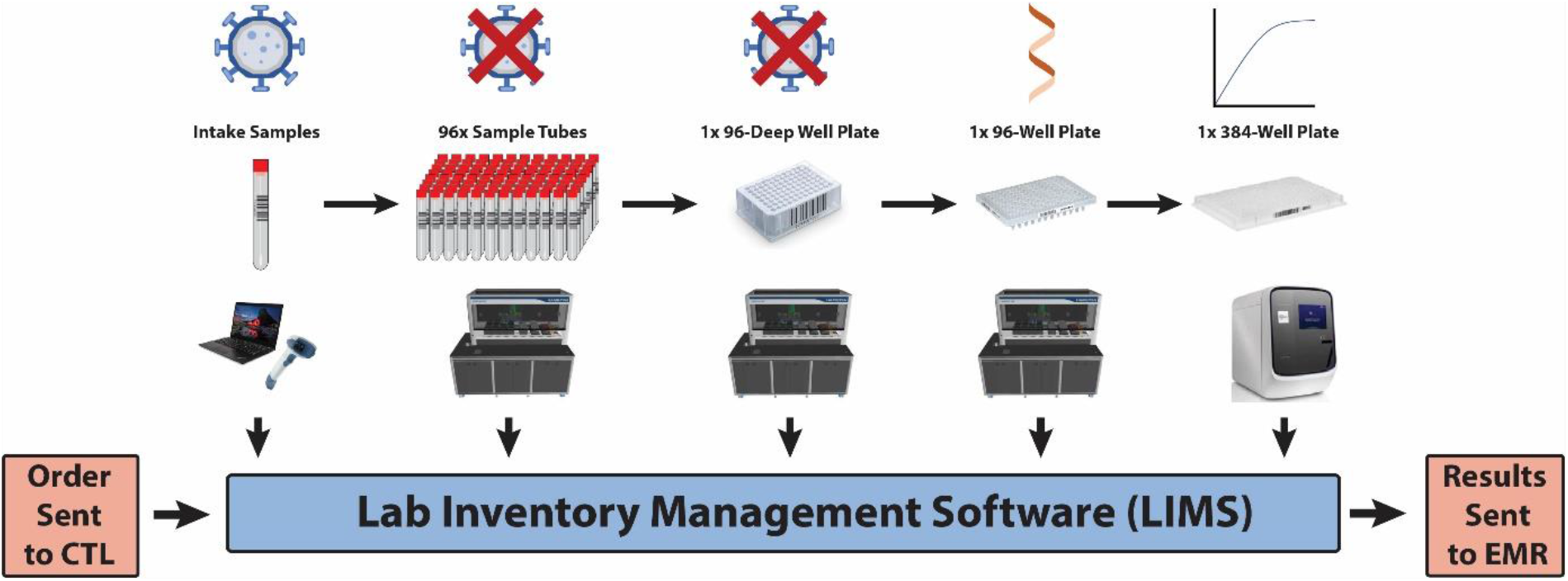
The two EMR systems send test orders to the CTL for each individual sample. The LIMS integration utilizes APIs to transfer information related to the sample during the laboratory’s testing process. This includes transferring information captured from each of the automation systems and the qPCR machine. The test results are reported out to the EMR systems through the LIMS.

To summarize a data workflow, each automation instrument takes in a data file and creates output files associated with barcoded plates and samples. These are tracked throughout the process up until results are exported from the completed qPCR plates. Each sample has a tracking lineage that includes lot numbers, sample process, plate information, and results. All results are checked by the Clinical Laboratory Supervisor prior to submission to the EMRs. All steps have duplicate manual checks to confirm results and sample integrity. Customization of the LIMS systems assured that each sample was fully back trackable to meet CLIA regulations.

### Legal and regulations

#### CLIA and MA Clinical Laboratory License

Before the pandemic, BU had held a long term CLIA license and corresponding Massachusetts Clinical Laboratory license for a high-complexity laboratory on the Charles River Campus. This small laboratory had been performing a genetic diagnostic test for a rare inborn error of metabolism called hereditary fructose intolerance^20^. Rather than applying for new CLIA and MA clinical laboratory licenses for the new COVID-19 laboratory, BU changed the location of its existing high-complexity laboratory’s licenses to the new on-campus COVID-19 laboratory. The construction of the CTL included a dedicated room for the continuation of genetic diagnostic testing inside the new laboratory. The co-location of the old laboratory and the new laboratory allowed BU to use the existing CLIA and MA clinical laboratory licenses to perform SARS-CoV-2 testing. The new laboratory was successfully inspected by MA Department of Public Health (DPH) in November 2020. The existence of this license and the long-standing relationship with the Massachusetts Department of Public Health greatly reduced the amount of paperwork required to move forward with test development and validation and allowed us to perform SARS-CoV-2 testing as soon as we completed validation testing without waiting for a MA DPH inspection. In addition, as the initial set up was nearing completion, BU hired a clinical laboratory supervisor with over 30 years of experience running CLIA laboratories.

#### FDA Emergency Use Authorization

In the spring and summer of 2020, the FDA was reviewing and granting emergency use authorization (EUA) status to laboratory developed tests (LDTs) like ours. We consulted with the FDA extensively and followed the template EUA for molecular LDTs^21^ during our validation testing. We submitted a pre-EUA document to the FDA on June 19, 2020. This submission put us in the queue for full review. We received previously tested discarded samples from collaborators at the Boston Medical Center, LabCorp Inc., Genova Diagnostics, and the Massachusetts Department of Public Health. Results of our successful validation experiments were compiled and submitted as a supplement to the pre-EUA on July 27, 2020. FDA regulations allow CLIA laboratories to run tests and deliver results once validation data is submitted and before the EUA is approved.

On August 19, 2020, the Department of Health and Human Services (HHS) under the Trump administration announced that the FDA may not require premarket review for LDTs, including EUA submissions, absent a notice-and-comment rulemaking process. HHS noted that laboratories may voluntarily submit an EUA request for LDTs if it desired to be eligible for coverage under the Public Readiness and Emergency Preparedness (PREP) Act, which provides certain liability protections for covered persons administering covered medical countermeasures. In response to the HHS announcement, the FDA in October 2020 announced that it would “declin[e] to review EUA requests for LDTs at this time,” including new EUA submissions and those already in the process of being reviewed. We continued (and continue) to maintain compliance with the FDA EUA requirements for our test, and HHS under the new Biden administration is still determining whether pending applications will be reviewed. CLIA requirements apply to clinical laboratories using LDTs, irrespective of a test’s EUA or approval status, and we continue to comply with all CLIA requirements.

## Results and discussion

### Limit of detection for SARS-CoV-2 plasmid

We documented the limit of detection (LoD) of the BU SARS-CoV-2 Test. A preliminary LoD study using IDT SARS-CoV-2 Plasmid Positive Control material spiked into the qPCR reaction was performed to assess the LoD with technical triplicates. The results showed a preliminary LoD of 10 copies per microliter (Table 1).

**Table 1.**
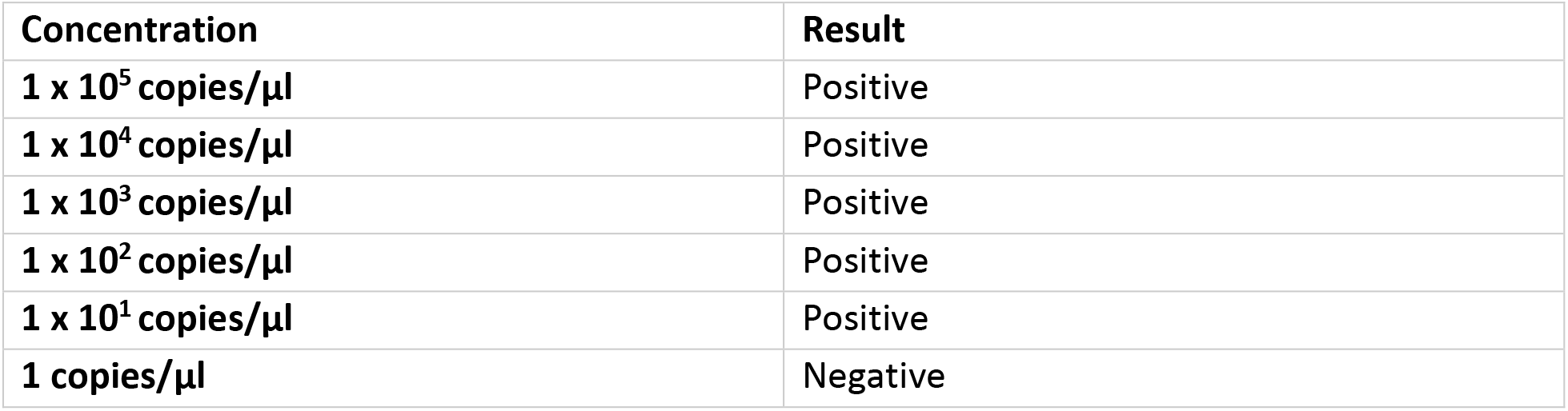
BU SARS-CoV-2 Test Preliminary LoD Study using IDT SARS-CoV-2 Plasmid Positive Control material. Preliminary LoD 10 copies per microliter.

| Concentration | Result |
| --- | --- |
| 1 x 10<sup>5</sup> copies/μl | Positive |
| 1 x 10<sup>4</sup> copies/μl | Positive |
| 1 x 10<sup>3</sup> copies/μl | Positive |
| 1 x 10<sup>2</sup> copies/μl | Positive |
| 1 x 10<sup>1</sup> copies/μl | Positive |
| 1 copies/μl | Negative |

Next, a known positive clinical specimen determined by an EUA-authorized test was used to generate dilutions in clinical matrix for LoD determination. Respiratory swab matrix solution from swab specimens collected from SARS-CoV-2 negative individuals was used as a diluent. We tested a 2-fold dilution series of three extraction replicates per concentration. The lowest concentration that gave positive results 100% of the time, 10.6 c/µl, was defined as the preliminary LoD (Table 2).

**Table 2:** BU SARS-CoV-2 Test LoD Study using Pooled Positive Residual Patient Samples. LoD 10.6 copies per microliter.

| Concentration | Result |
| --- | --- |
| 85 copies/μl | Positive |
| 42.5 copies/μl | Positive |
| 21.3 copies/μl | Positive |
| 10.6 copies/μl | Positive |
| 5.3 copies/μl | Negative |
| 2.7 copies/μl | Negative |

The final LoD concentration was be confirmed by testing 20 individual extraction replicates at the preliminary LoD. The LoD, 10.6 c/µl, was the lowest concentration at which 20/20 replicates were positive (Table 3).

**Table 3:**
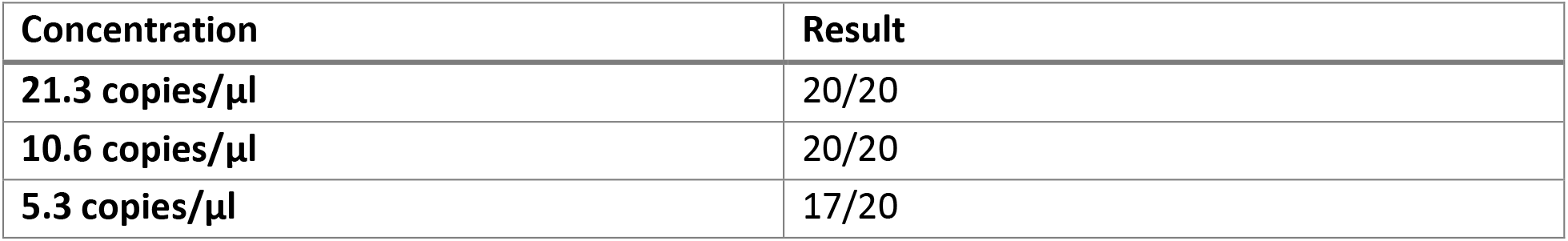
BU SARS-CoV-2 Test LoD Confirmation Study using replicates of 20 Positive Residual Patient Samples. LoD established at 10.6 copies per microliter.

### Modifying existing testing for pooled sample testing

The development and distribution of vaccines in the US has resulted in a decrease in Covid-19 cases and deaths, which leads to the feasibility of implementing pooled sample testing^22,23^. The BU CTL planned and implemented pooled testing for the university. In a 1-month period during the Summer of 2021, the workflow for individual sample testing was modified by implementing algorithmic changes in the script responsible for sample data handling and analysis. In addition, the changes were implemented with updates to the Accession and Inactivation steps to pool 5 individual samples prior to the automation steps. Pooling has allowed for an increase in sample throughput while reducing the total reagent and consumable use.

### Considerations and future research opportunities

BU has successfully implemented a screening testing program for the 2020-2021 academic school year with the plan to continue to allow for students, staff, and faculty to safely return to campus. With over 1 million tests completed this academic year, the university has the unique opportunity for research to contribute to the SARS-CoV-2 knowledgebase. The university has a controlled data diverse set of de-identified samples that could provide further insight into SARS-CoV-2 and its impacts on public health.

## Data Availability

Data are available upon request.

## Acknowledgements

Figures were drawn by the Authors of this paper. Images are stock from ThermoFisher, Hamilton, Zebra, SHI, LabConco, and Boston University.

The authors of the paper would like to thank our collaborators from across the country including the Broad Institute, University of California Berkeley, and University of California San Diego.

Thank you to Sandra Smole and Massachusetts Department of Public Health for their continued support.

Thank you to Marcia Eisenberg and LabCorp, Inc. for technical assistance and advice.

Thank you to Amy Peace Brewer and Genova Diagnostics for technical assistance and advice.

Thank you to the support of Rajen Kilachand and the Rajen Kilachand Center for Integrated Life Sciences & Engineering for the support to develop the BU CTL.

Thank you to the technicians, technologists, and Lab Managers of the BU CTL. The authors would like to thank the BU Facilities Management and Operations.

